# Olanzapine for the Prevention and Treatment of Nausea and Vomiting Induced by Chemotherapy for Lung Cancer: protocol for a multicenter, double-blind and randomized controlled trial

**DOI:** 10.1101/19012583

**Authors:** Jian-Guo Zhou, Pei-Jie Li, Su-Han Jin, Da-Hai Liu, Lang Huang, Ming-Ze Cao, Yu Zou, Hu Ma

**Author notes:** Correspondence to: Jian-Guo Zhou MD, Department of Oncology, Affiliated Hospital of Zunyi Medical University, Zunyi, China, NO.149, Dalian Road, Zunyi 563000, China. Hu Ma MD PhD, Department of Oncology, Affiliated Hospital of Zunyi Medical University, Zunyi, China, NO.149, Dalian Road, Zunyi 563000, China.; Email: l. Jian-Guo Zhou and Pei-Jie Li contributed equally to this work.

## Abstract

**Background:** Chemotherapy-induced nausea and vomiting (CINV) is frequently observed after the administration of chemotherapy and significantly influences the quality of life (QoL) of patients. Olanzapine has a high control rate of CINV in patients with cancer when combined with the NK-1 receptor antagonist dexamethasone and 5-hydroxytryptamine (5-HT3) receptor antagonists. The efficacy of a regimen without an NK-1 receptor antagonist remains unknown. Therefore, we designed this randomized trial to provide evidence for the management of CINV.

**Methods and ananlysis:** This is a double-blind, multicenter and randomized controlled trial. Patients with histologically confirmed lung cancer will be assessed by physicians based on the inclusion and exclusion criteria, and 156 participants will be enrolled and randomized to a placebo group or experiment group to receive treatment for CINV. The primary endpoint is the incidence of delayed CINV. The secondary endpoints are complete response (CR) of acute CINV, CR of delayed CINV, effective control rate (ECR) of CINV and QoL. During the six days after administration, these endpoints will be evaluated and recorded by physicians.

**Ethic and dissemination:** This study has received approval from the institutional ethical review board of the Affiliated Hospital of Zunyi Medical University (ref approval No. 58). Written informed consent will be signed by all participants prior to enrolling. Participants will be randomly assigned to the experimental group or comparator group by blocked randomization.

**Article summary:** Strengths and limitations

- This study will provide evidence for physicians to find an affordable and effective treatment regimen for CINV through this study.
- Because of the variety of pharmaceutical companies, medical care and other factors, problem of cost will be further explored in the following studies.
- Although the study is designed as a double-blind randomized controlled trial, QoL will be measured by a questionnaire that is filled out by patients themselves. This may influence the conclusions.

## Background

Chemotherapy-induced nausea and vomiting (CINV) is frequently observed in the treatment of cancer, and influences the quality of life (QoL) of patients and their adherence to treatment. CINV also leads to dehydration, malnutrition and other adverse events^[1,2]^. Therefore, the prevention and relief of CINV are indispensable to ensure the conduction of chemotherapy. The mechanism of CINV remains unknown, and most studies have shown that CINV is mainly related to the following aspects. First, chemotherapeutic agents stimulate the gastrointestinal tract, which induces the release of neurotransmitters by chromaffin cells. The neurotransmitters bind to their corresponding receptors, which then results in vomiting by stimulating the vomiting center. Second, Chemotherapeutic agents and their metabolites directly activate chemoreceptor. Third, mental factors directly irritate the cerebral cortex pathway^[3]^. The major neurotransmitters that lead to vomiting include dopamine (DA), histamine, 5-hydroxytryptamine (5-HT), and substance P^[4-6]^. Studies have demonstrated that 5-HT is related to acute CINV. Thus, 5-HT3 receptor antagonists, such as granisetron, could be an effective medicine for CINV. Geling and colleagues declared that a 5-HT3 receptor antagonist was effective for acute CINV but had little efficacy in preventing delayed emesis^[7-8]^. In addition, studies have indicated that the NK-1 receptor antagonist aprepitant is a potent agent in relieving CINV. Four phase 3 trials indicated that aprepitant had a significantly higher control rate of CINV in patients with cancer than standard antiemetic therapy, which made aprepitant indispensable in the management of emetogenic chemotherapy^[9-13]^. Thus, correlative guidelines recommend regimens with a 5-HT3 receptor antagonist, NK-1 receptor antagonist and glucocorticoid as the standard treatment for strongly emetic chemotherapy regimens^[8]^. However, the prevention of nausea and vomiting caused by moderately emetic chemotherapy regimens remains a problem in clinical practice.

Olanzapine inhibits several neurotransmitters that cause CINV, which is why this medicine is effective for both acute and delayed CINV^[14]^. Olanzapine is widely used in the treatment of mental disorders. Thus, this drug can alleviate anxiety, improve sleep quality and relieve pain from opioid therapy in patients with cancer^[15]^. The usual adverse effects of olanzapine are lethargy, weight gain, fatigue, constipation, hyperlipidemia and hyperglycemia. The most common adverse effect is lethargy, which can oppose the insomnia and excitation caused by dexamethasone^[16]^. In summary, olanzapine is a potent agent to control CINV with mild adverse effects, and the use of olanzapine is worth generalizing. However, there are problems to need to be solved in the application of olanzapine for CINV. Aprepitant, which is included in the recommended treatment method of CINV, is expensive. This limits its application in patients. Additionally, no clinical trial has been designed to prove the efficacy of olanzapine in antiemetic therapy when it is not in combination with an NK-1 receptor antagonist.

Because of these reasons, we intend to compare the regimens with olanzapine (or a placebo), dexamethasone and 5-HT3 receptor antagonists in terms of efficacy and adverse events during the treatment of CINV. We aim to provide an available therapeutic option for CINV in patients with cancer to improve their QoL.

## Methods

### Study design

This study is a multicenter, double-blind and randomized controlled trial. Olanzapine will be compared with a placebo (both are combined with dexamethasone and 5-HT3 receptor antagonists) to determine if olanzapine is an effective and safe choice for patients suffering from CINV. We plan to include patients with lung cancer regardless of their histopathologic types and sex, and a subgroup analysis will be conducted later to estimate whether these differences influence the efficacy of olanzapine. A total of 156 subjects will be randomized into two groups (experimental group or comparator group). This study will be conducted in the Affiliated Hospital of Zunyi Medical University, Zunyi, China; Sichuan Cancer Hospital and Research Institute, Chengdu, China; Affiliated Hospital of North Sichuan Medical College, Nanchong, China; First People’s Hospital of Zunyi, Zunyi, China; Affiliated Hospital of Southwest Medical University, Luzhou, China; and Guizhou Provincial People’s Hospital, Guiyang, China. The procedures of this study are shown in *Fig. 1*.

**Figure 1:**
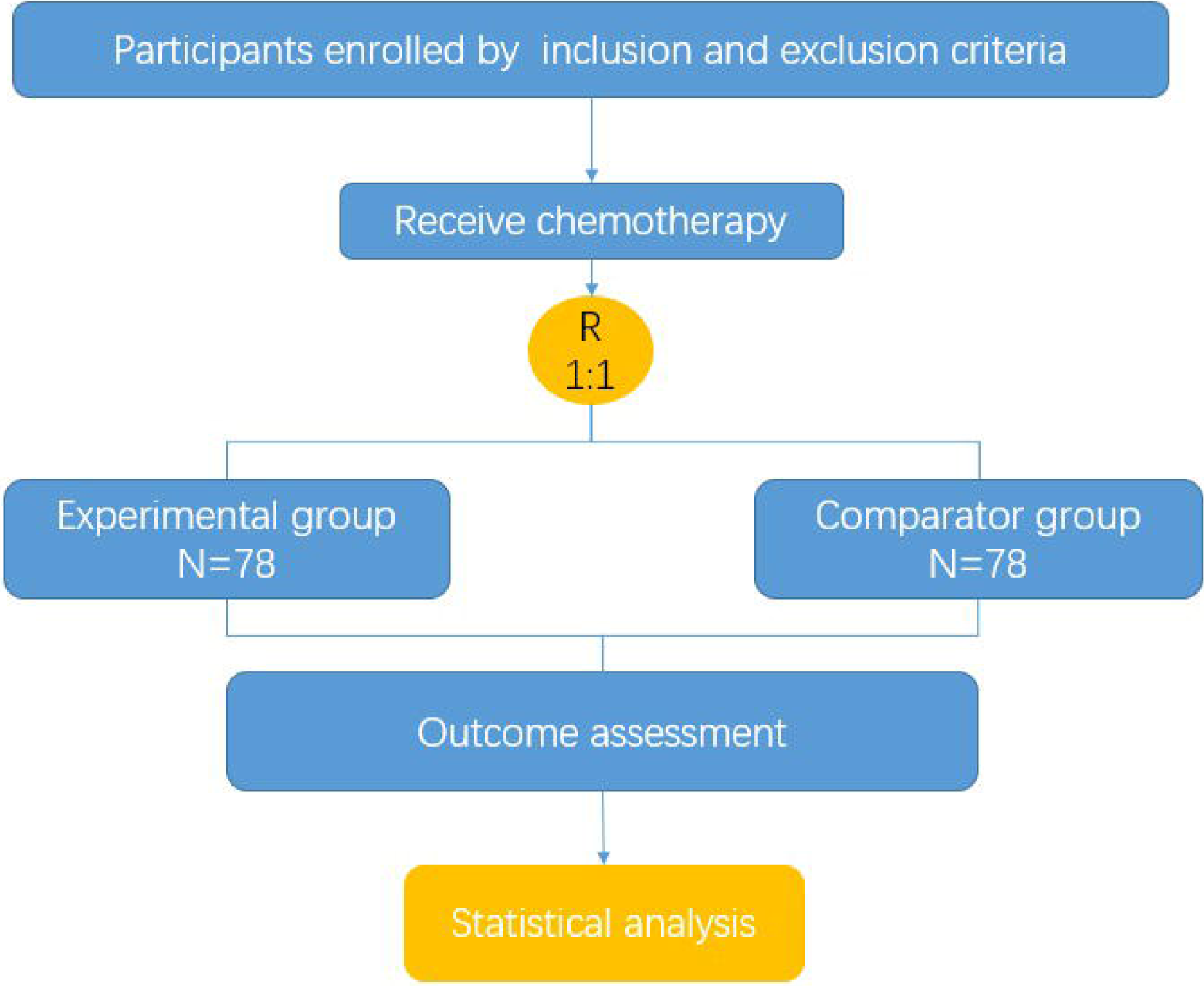

### Inclusion criteria

1. Patients aged 18 to 70 years, male or female.
2. Patients with an Eastern Cooperative Oncology Group (ECOG) performance status ≤ 2 or Karnofsky performance status (KPS) scores ≥ 60.
3. Patients with cytologically or histologically confirmed lung cancer (including adenocarcinoma, squamous carcinoma, small-cell lung cancer and other types of lung cancers).
4. Patients who agree to receive chemotherapy and who can tolerate at least 2 cycles of chemotherapy (no matter how many cycles of chemotherapy the patient previously received).
5. Chemotherapy regimens recommended by the clinical practice guidelines (National Comprehensive Cancer Network guidelines and Chinese Society of Clinical Oncology guidelines of lung cancer). Specific treatments include cisplatin or carboplatin (AUC≥4) plus pemetrexed, cisplatin or carboplatin (AUC≥4) plus docetaxel, cisplatin or carboplatin (AUC≥4) plus paclitaxel and cisplatin or carboplatin (AUC≥4) plus etoposide. Specific dosages will be calculated according to the body surface area of the participants. The dosage of cisplatin can be divided over three days for administration. The duration of highly emetic therapy should be less than or equal to three days.
6. Patients with adequate organ function including the following: Adequate bone marrow reserve: white blood cell (WBC) count higher than or equal to 2.0 ×10^9^/L, absolute neutrophil count (ANC) superior or equal to 1.5 ×10^9^/L, platelets higher than superior or equal to 80 X 10^9^/L, and hemoglobin higher than or equal to 90g/L. Hepatic parameters: bilirubin <1.5 times the upper limit of normal (ULN) and aspartate transaminase (AST) and alanine transaminase (ALT) ≤2.5xULN (or <5xULN with liver metastases); and Renal parameters: Serum creatinine≤1×ULN, calculated creatinine clearance (CrCl) higher than or equal to 50mL/min based on the standard Cockroft and Gault formula.
7. There should be at least 3 weeks from the end of last chemotherapy.
8. Women of reproductive years who are willing to avoid pregnancy with the appropriate methods during the study and 8 weeks after the last administration. A pregnancy test before beginning the administration will be conducted when necessary, and the results need to be negative.
9. Participants who are willing to join this study and sign the informed consent form.

### Exclusion criteria

Patients with any one of these following criteria will be excluded:

1. Women who are pregnant or breastfeeding.
2. Patients who need to receive radiotherapy during this trial.
3. Patients with an obstruction of the gastrointestinal tract.
4. Patients with severe heart, renal or liver disease.
5. Patients with metabolic abnormalities.
6. Patients with epilepsy or who are using sedatives.
7. Patients who have received an with antiemetic less than 24 hours before chemotherapy or those who have suffered from vomiting before chemotherapy.
8. Patients with brain metastases (intracranial hypertension may cause vomiting).
9. Patients with contraindications to chemotherapy.
10. Patients who have an allergic reaction to treatment in this study.
11. Patients who are participating in another clinical trial or who have participated in a study less than 2 weeks prior.
12. Patients who are considered unsuitable to be included by the treating physicians.

#### Patient and public involvement

There will be two groups in this study (experimental group and comparator group). All participants will received dexamethasone (10 mg/d, from the first day to the third day) plus tropisetron (4 mg/d, from the first day to the third day).The patients in the comparator group will receive a placebo. The patients in the experimental group will receive olanzapine (10 mg/d). Placebo and olanzapine will be given from the first day to the fourth day. Both of them have the same packaging, and their quality has been tested by Jiangsu Hansoh Pharmaceutical Co., Ltd according to China’s State Food and Drug Administration standards YBH05472018 (No.FC190082). The intervention will provide for only one cycle. The time schedule of the enrollment, interventions and assessments of this study is shown in Table 2. First of all, patients will be informed by researchers or physicians about the research questions and the outcome measurements. An informed consent will be signed by all participants prior to enrolling. Participants will be randomly assigned (1:1) to the experimental group or comparator group by blocked randomization, and this procedure will be stratified by center. We will use the “Statistical Product and Service Solutions” (SPSS) software to generate the random number table. Each center will designate a specific staff member who is responsible for the storage, distribution, recording and recycling of the medicine. All bags which contain Olanzapine or placebo will be numbered from 1 to 156 according to the random number table. And patients will then finished their own questionnaires to evaluated the QoL. Patients and physicians will not be aware of which strategy is selected. Researchers are not blinded to the randomization, so they are responsible for measuring the outcomes and analyzing the data. The results of study will disseminate to participants by researchers through phone or e-mail after publishing.

### Outcome measures

#### Primary endpoint

Incidence of delayed CINV (CINV that occurred from 24 hours to 120 hours after chemotherapy).

#### Secondary endpoint

Complete response (CR) of acute CINV, CR of delayed CINV, effective control rate (ECR) (CR plus partial response), and QoL.

Because of the difficulty to include enough participants who did not receive chemotherapy before, all assessments are conducted after any cycle of chemotherapy. In terms of the influence of cycles of chemotherapy on the frequency of events, we will conduct subgroup to elucidate this problem.

### QoL assessment

The QoL of the participants will be measured by the European Organization for Research and Treatment of Cancer Quality of Life Questionnaire-Lung Cancer 13 (EORTC QLQ-LC13) scale, the European Organization for Research and Treatment of Cancer Quality of Life (EORTC QLQ-LC30) scale and the Functional Assessment of Cancer Therapy-Lung cancer (FACT-L) scale ^[17-19].^ The EROTC QLQ-LC13 and EORTC QLQ-LC30 scales comprise several symptoms measured by scores ranging from 1 to 7; the higher the symptoms are scored, the worse the QoL is. The FACT-L scale is a combination of the 27-item FACT-General (FACT-G) scale and the 9-item Lung Cancer Subscale (LCS) and measures multidimensional quality of life. The higher the score obtained, the better the QoL is. The use of all scales has been authorized.

### Safety evaluation

The safety of olanzapine will be assessed by the following aspects: vital sign measurements, routine blood examinations, liver and renal function tests, electrocardiographs and the occurrence of relevant adverse events (AEs). AEs are unintended situations that occur after the use of experimental therapy, and not all of AEs result from treatment. AEs are divided into mild (treatment is usually not needed), moderate (moderate damage to the body) and severe AEs (severe damage to the body and is life-threatening) according to symptoms of the participants and degree of injury to the organs caused by the drugs. Investigators will also evaluate the degree of relevance between the AEs and the experimental therapy. The time of appearance, severity level, duration, relevant treatment measurements and outcomes of the AEs will be recorded by the researchers. A serious adverse event (SAE) is an adverse reaction that meets one of the following criteria: 1) causes death; 2) causes a high risk of death; 3) results in hospitalization or a longer hospital stay; 4) leads to permanent or severe disability. When AEs are observed in the participants, investigators can terminate the treatment according to the severity of the AEs and give corresponding therapy.

### Statistical methods

We will use SPSS software to conduct statistical analysis. The actual number of participants, the number of participants lost, the number of excluded cases, basic characteristics, patient compliance and efficacy rates and safety of olanzapine will be analyzed. Mean± standard deviation will be used to describe quantitative data that are normally distributed. The interquartile range (IQR) will be used to describe quantitative data that are not normally distributed. We will use the t-test or Wilcoxon rank-sum test to analyze quantitative data. The chi-square test will be used to analyze the count data. A P value of less than 0.05 woll be regarded as statistically significant. All the data in this study are based on the Electronic Data Capture (EDC) system, and the system is developed and supported by Jiangsu famisheng medical Co. Ltd. The data administrator will establish an electronic medical record report (eCRF) and is responsible for the confirmation from the monitor, applicant and principal investigators. The system administrators are responsible for the confirmation from the data administrators.

### Sample size

The determination of sample size refers to the schemes and results of studies conducted by Navari[20] Studies have shown that the incidence of delayed CINV in patients who receive olanzapine plus standard treatment is 68-83% and in patients who only received standard treatment is 23-58%. Therefore, we chose 68% for the experimental group and 40% for the comparator group. We chose 0.05 as the type I error rate, and the type II rate was 0.0997 by calculation. With the use of PASS 11.0, each group needs 62 participants. We estimate that 20% of the participants will withdraw from the study or be lost to follow-up and thus, each group needs 78 participants.

### Data monitoring

Investigators and physicians will strictly follow the study protocol, and all procedures will be standard and professional. There is minimal risk of harm with olanzapine for the treatment of CINV according to the reported study, and all the adverse events caused by olanzapine will be evaluated as secondary outcomes. Therefore, a Data Monitoring Committee (DMC) is not needed.

## DISCUSSION

Patients with lung cancer have sufferd from CINV for decades, which influences their QoL to a large extent. Clinical trials have demonstrated that aprepitant has a significantly higher control rate of CINV in patients with cancer compared with standard antiemetic therapy^[9-13]^. Regimens with a 5-HT3 receptor, NK-1 receptor antagonist and glucocorticoid have been recommended for CINV in the clinical guidelines. However, in China, the NK-1 receptor antagonist aprepitant is not covered in medical care, and many patients cannot afford this medicine.

Olanzapine is efficient in the treatment of CINV according to several clinical studies, although this drug mainly used in therapy for mental disorders^[18-20]^. A systematic review elucidated that olanzapine regimens achieved a higher complete response in the overall and delayed phases than traditional antiemetic drugs^[21]^. Since there are no large clinical trials that explicitly investigate the efficacy and safety of olanzapine not in combination with NK-1 receptor antagonists in CINV, guidelines in China do not recommend olanzapine to alleviate nausea and vomiting. Furthermore, 5-HT3 receptor antagonists were administered by intravenous injection (iv), while in our study, the drugs were administered by intravenously guttae (ivgtt). Dexamethasone was used by orally in most studies, but in our study, the drug was administered ivgtt^[20-22]^. Therefore, participants will stay in the hospital during this study, which allows for the evaluation to be highly detailed and correct. We hope to find an affordable and effective treatment regimen for CINV through this study.

Because of the variety of pharmaceutical companies, medical care and other factors, we will not discuss the problem of cost in this study. Data will be collected, and this problem will be further explored in the following studies. Although the study is designed as a double-blind randomized controlled trial, QoL will be measured by a questionnaire that is filled out by patients themselves. This may influence the conclusions of this study.

## Trial Status

This study has been registered on www.clinicaltrials.gov (ID: NCT03571126), and recruitment is expected to begin in June, 2019.

## Data Availability

All of these study data are available upon reasonable request and can be obtained by contacting with the corresponding author.

## Declarations

### Ethics approval and consent to participate

This study has received approval from the institutional ethical review board of the Affiliated Hospital of Zunyi Medical University (ref approval No. 58) and we will not begin recruiting at subcenters in the trial until local ethical approval has been obtained. Subjects will obtain compensation if SAE occurs. The details are listed in the policy schedule drafted by Asia-Pacific Property & Casualty Insurance Co., Ltd (NO.06020400000912362019000024).

Written informed consent (see Additional file 1) will be signed by all participants prior to enrolling, PI or clinical researchers will inform participants about the procedures of this study, what regimen will be administrated and so on. Patients will have enough time to considerate whether they are willing to join this trial. The patients will receive a copy of this form. Withdrawal from the study is allowed at any time if the patient is unwilling to continue the trial. After withdrawal of the study, data from these patients will not be included in the data analysis.

### Authors’ contributions

JG Zhou MD is the first author who designed this study and also drafted, read, revised and approved the final version of this manuscript. PJ Li MM is the co-first author who contributed equally to this work. JG Zhou MD and Prof. Hu Ma are the corresponding authors who conceived and designed the study, modified and read and approved the final version. JG Zhou, PJ Li, DH Liu, SH Jin, MZ Cao and Y Zou are responsible for patient recruitment, assign participants to interventions and follow up.

### Authors’ Information

#### Protocol contributors

The main information of authors of this protocol is listed in *Table 1*.

**Table 1.**
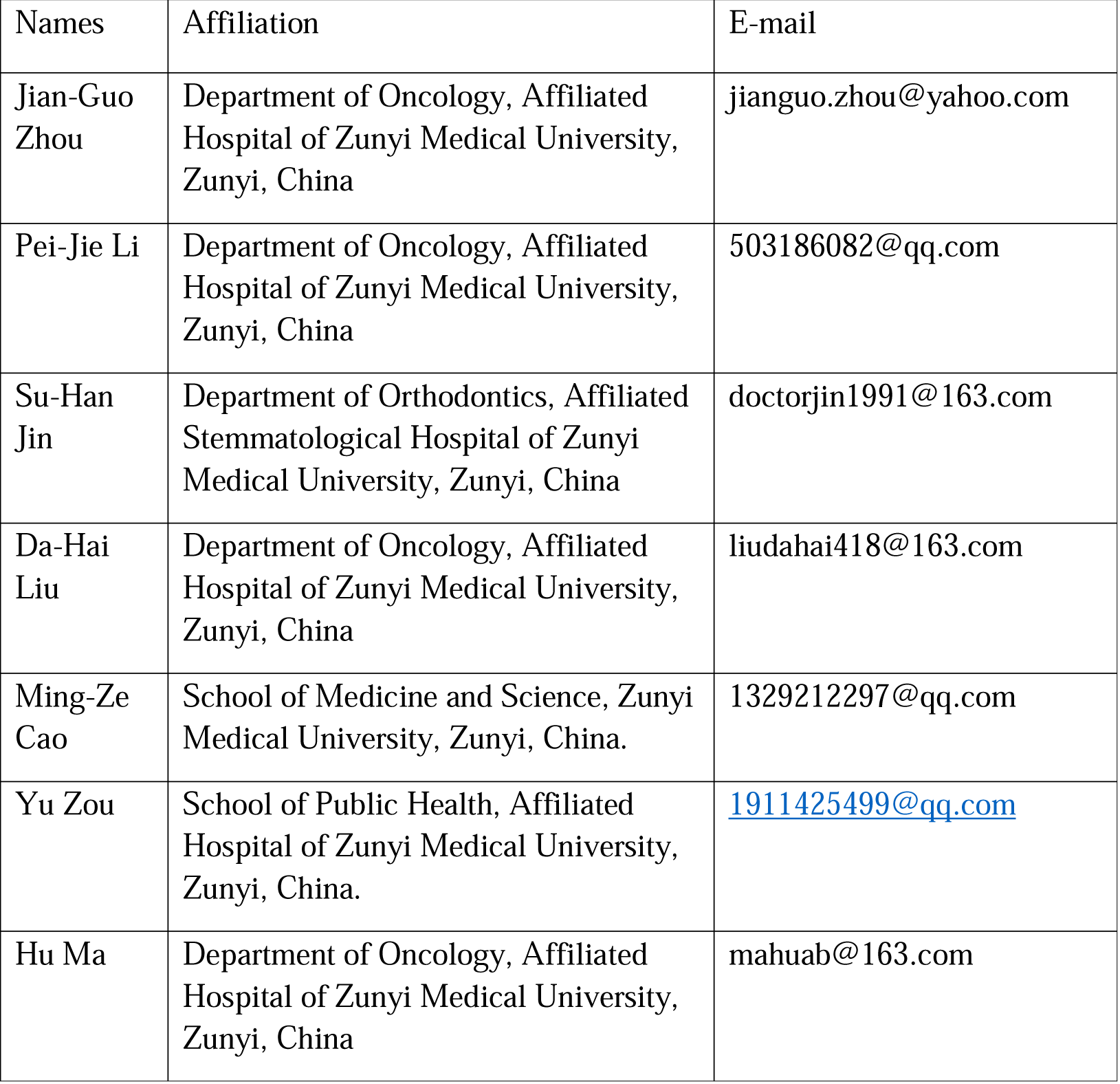
The main information of the authors

**Table 2.**
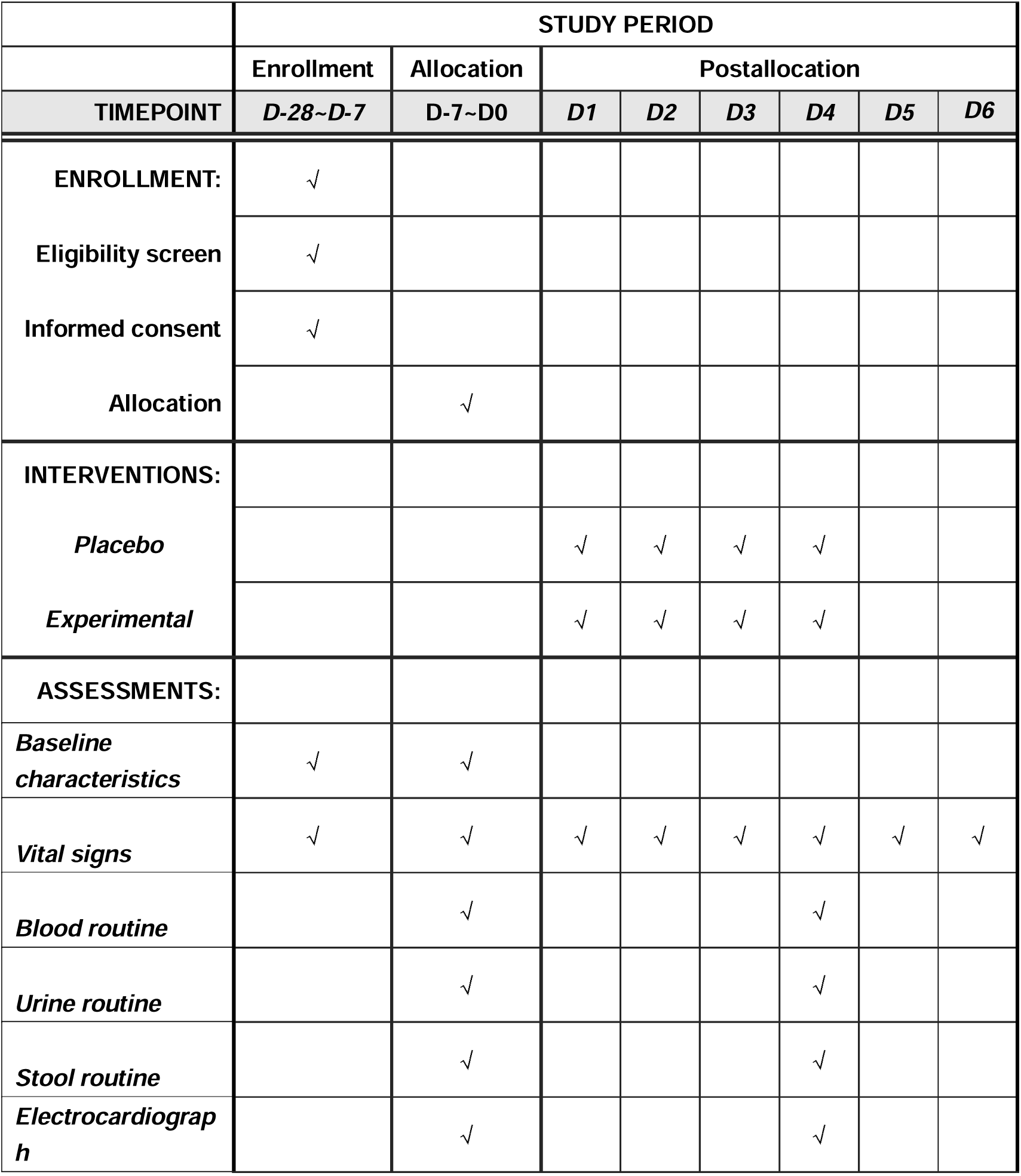
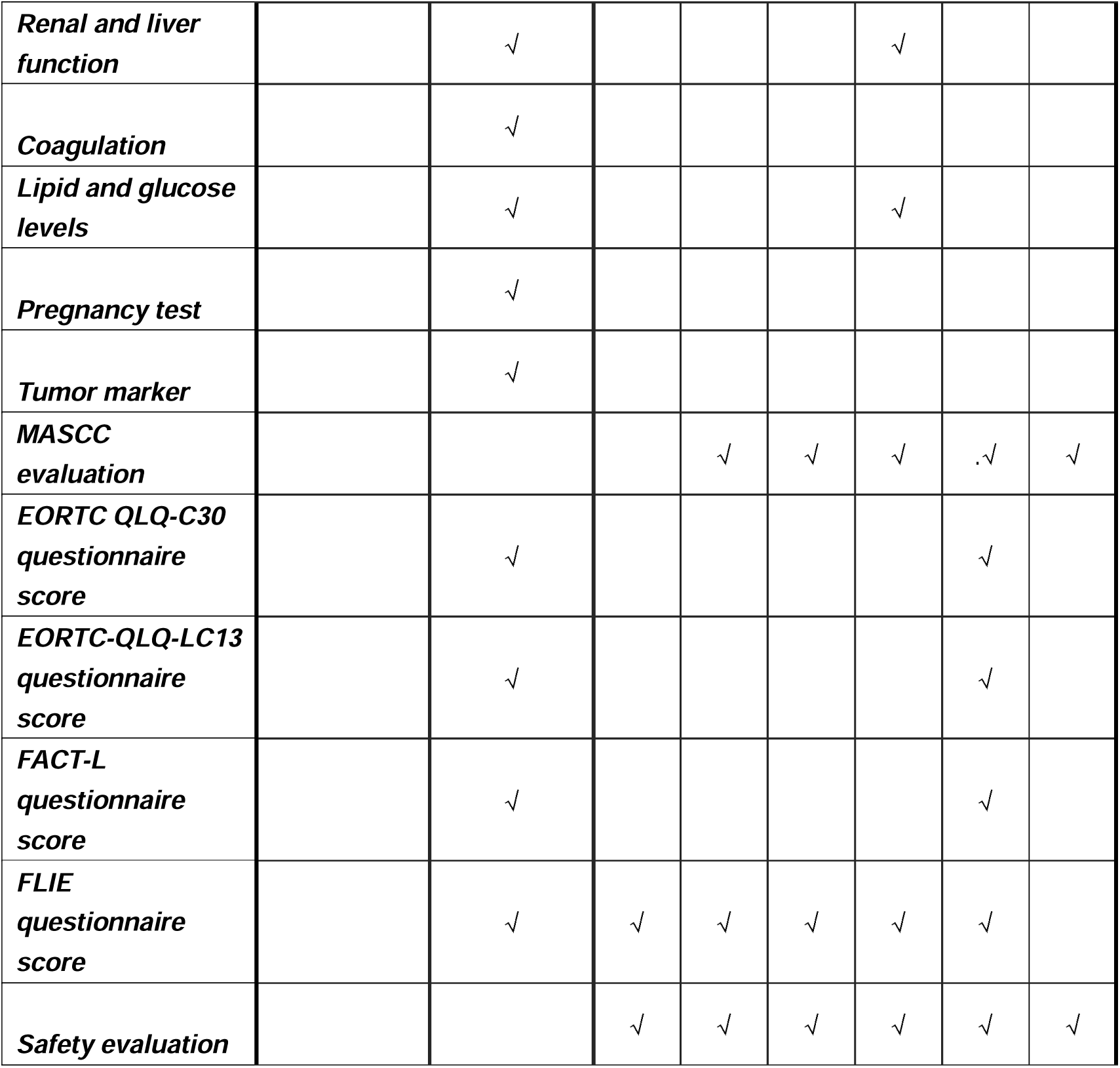
Time schedule of the enrollment, interventions, and assessments

|  | STUDY PERIOD |  |  |  |  |  |  |  |
| --- | --- | --- | --- | --- | --- | --- | --- | --- |
|  | Enrollment | Allocation | Postallocation |  |  |  |  |  |
| TIMEPOINT | D-28~D-7 | D-7~D0 | D1 | D2 | D3 | D4 | D5 | D6 |
| **ENROLLMENT:** | √ |  |  |  |  |  |  |  |
| Eligibility screen | √ |  |  |  |  |  |  |  |
| Informed consent | √ |  |  |  |  |  |  |  |
| Allocation |  | √ |  |  |  |  |  |  |
| **INTERVENTIONS:** |  |  |  |  |  |  |  |  |
| Placebo |  |  | √ | √ | √ | √ |  |  |
| Experimental |  |  | √ | √ | √ | √ |  |  |
| **ASSESSMENTS:** |  |  |  |  |  |  |  |  |
| Baseline characteristics | √ | √ |  |  |  |  |  |  |
| Vital signs | √ | √ | √ | √ | √ | √ | √ | √ |
| Blood routine |  | √ |  |  |  | √ |  |  |
| Urine routine |  | √ |  |  |  | √ |  |  |
| Stool routine |  | √ |  |  |  | √ |  |  |
| Electrocardiograph |  | √ |  |  |  | √ |  |  |
| **Renal and liver function** |  | √ |  |  |  | √ |  |  |
| **Coagulation** |  | √ |  |  |  |  |  |  |
| **Lipid and glucose levels** |  | √ |  |  |  | √ |  |  |
| **Pregnancy test** |  | √ |  |  |  |  |  |  |
| **Tumor marker** |  | √ |  |  |  |  |  |  |
| **MASCC evaluation** |  |  |  | √ | √ | √ | .√ | √ |
| **EORTC QLQ-C30 questionnaire score** |  | √ |  |  |  |  | √ |  |
| **EORTC-QLQ-LC13 questionnaire score** |  | √ |  |  |  |  | √ |  |
| **FACT-L questionnaire score** |  | √ |  |  |  |  | √ |  |
| **FLIE questionnaire score** |  | √ | √ | √ | √ | √ | √ |  |
| **Safety evaluation** |  |  | √ | √ | √ | √ | √ | √ |

**Table 3.**
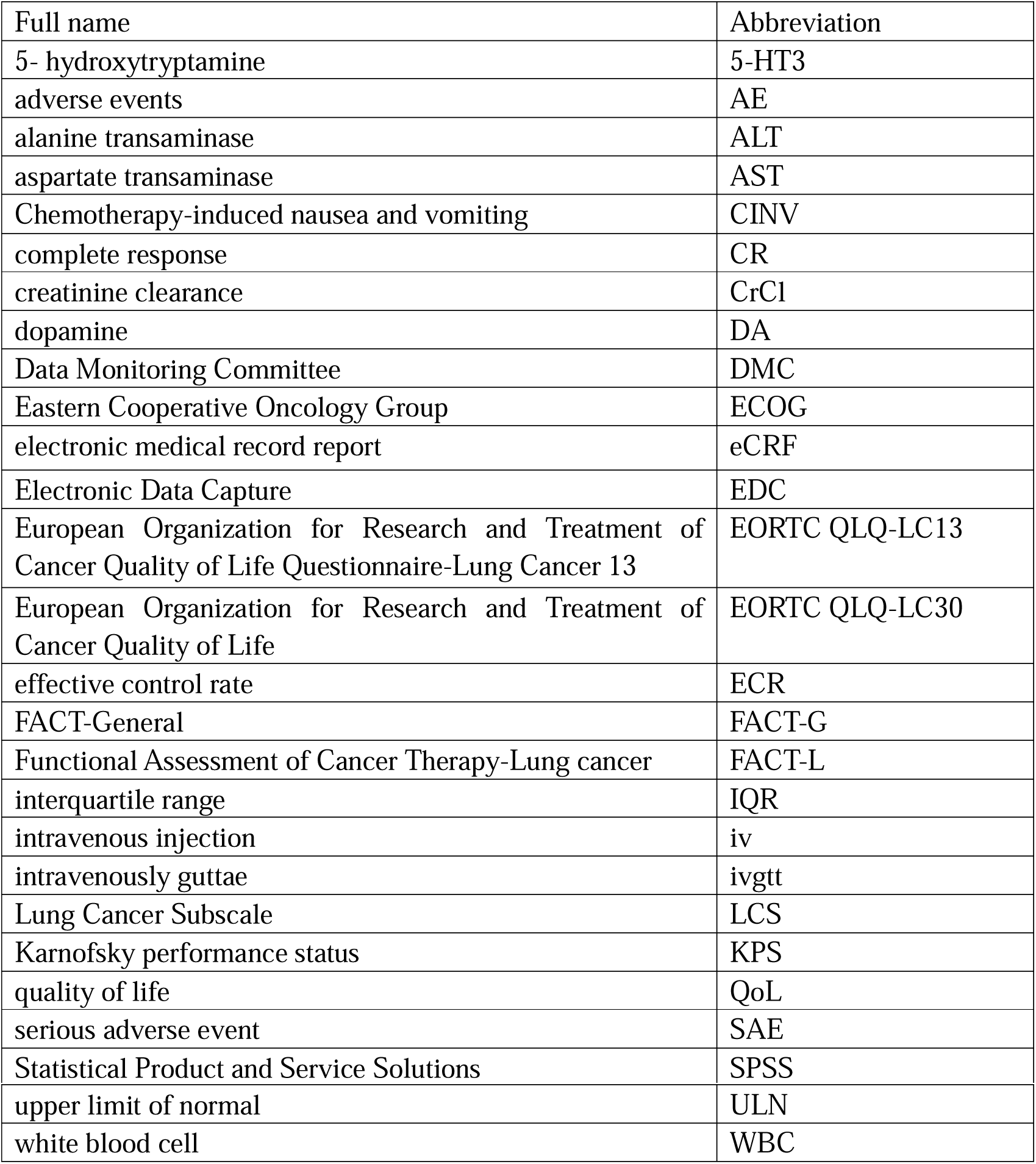
List of abbreviations

| Full name | Abbreviation |
| --- | --- |
| 5- hydroxytryptamine | 5-HT3 |
| adverse events | AE |
| alanine transaminase | ALT |
| aspartate transaminase | AST |
| Chemotherapy-induced nausea and vomiting | CINV |
| complete response | CR |
| creatinine clearance | CrCl |
| dopamine | DA |
| Data Monitoring Committee | DMC |
| Eastern Cooperative Oncology Group | ECOG |
| electronic medical record report | eCRF |
| Electronic Data Capture | EDC |
| European Organization for Research and Treatment of Cancer Quality of Life Questionnaire-Lung Cancer 13 | EORTC QLQ-LC13 |
| European Organization for Research and Treatment of Cancer Quality of Life | EORTC QLQ-LC30 |
| effective control rate | ECR |
| FACT-General | FACT-G |
| Functional Assessment of Cancer Therapy-Lung cancer | FACT-L |
| interquartile range | IQR |
| intravenous injection | iv |
| intravenously guttae | ivgtt |
| Lung Cancer Subscale | LCS |
| Karnofsky performance status | KPS |
| quality of life | QoL |
| serious adverse event | SAE |
| Statistical Product and Service Solutions | SPSS |
| upper limit of normal | ULN |
| white blood cell | WBC |

#### Corresponding authors

Correspondence to: Jian-Guo Zhou MD, Department of Oncology, Affiliated Hospital of Zunyi Medical University, Zunyi, China, NO.149, Dalian Road, Zunyi 563000, China. Tel: +86-0851-28609038; Fax: +86-0851-28609095. Hu Ma MD PhD, Department of Oncology, Affiliated Hospital of Zunyi Medical University, Zunyi, China, NO.149, Dalian Road, Zunyi 563000, China. Tel: +86-0851-28609038; Fax: +86-0851-28609095; Email: l.

## Acknowledgements

We thank Sichuan Cancer Hospital and Research Institute, Chengdu, China; Affiliated Hospital of North Sichuan Medical College, Nanchong, China; The First People’s Hospital of Zunyi, Zunyi, China; The affiliated hospital of southwest medical university, Luzhou, China; and Guizhou Provincial People’s Hospital, Guiyang, China for their collaboration with us.

## Abbreviations

All the abbreviations used in this protocol has been listed in Table 3.

